# Impact of Social Vulnerability on COVID-19 Incidence and Outcomes in the United States

**DOI:** 10.1101/2020.04.10.20060962

**Authors:** Aditi Nayak, Shabatun J. Islam, Anurag Mehta, Yi-An Ko, Shivani A. Patel, Abhinav Goyal, Samaah Sullivan, Tené T. Lewis, Viola Vaccarino, Alanna A. Morris, Arshed A. Quyyumi

## Abstract

**Importance:** Prior pandemics have disparately affected socially vulnerable communities. Whether regional variations in social vulnerability to disasters influence COVID-19 outcomes and incidence in the U.S. is unknown.

**Objective:** To examine the association of Social Vulnerability Index (SVI), a percentile-based measure of county-level social vulnerability to disasters, and its sub-components (socioeconomic status, household composition, minority status, and housing type/transportation accessibility) with the case fatality rate (CFR) and incidence of COVID-19.

**Design:** Ecological study of counties with at least 50 confirmed COVID-19 cases as of April 4^th^, 2020. Generalized linear mixed-effects models with state-level clustering were applied to estimate county-level associations of overall SVI and its sub-component scores with COVID-19 CFR (deaths/100 cases) and incidence (cases/1000 population), adjusting for population percentage aged ≥65 years, and for comorbidities using the average Hierarchical Condition Category (HCC) score. Counties with high SVI (≥median) and high CFR (≥median) were identified.

**Setting:** Population-based study of U.S. county-level data.

**Participants:** U.S. counties with at least 50 confirmed COVID-19 cases.

**Main outcomes and measures:** COVID-19 CFR and incidence.

**Results:** Data from 433 counties including 283,256 cases and 6,644 deaths were analyzed. Median SVI was 0.46 [Range: 0.01-1.00], and median CFR and incidence were 1.9% [Range: 0-13.3] and 1.2 per 1000 people [Range: 0.6-38.8], respectively. Higher SVI, indicative of greater social vulnerability, was associated with higher CFR (RR: 1.19 [1.05, 1.34], p=0.005, per-1 unit increase), an association that strengthened after adjustment for age≥65 years and comorbidities (RR: 1.63 [1.38, 1.91], p<0.001), and was further confirmed in a sensitivity analysis limited to six states with the highest testing levels. Although the association between overall SVI and COVID-19 incidence was not significant, the SVI sub-components of socioeconomic status and minority status were both predictors of higher incidence and CFR. A combination of high SVI (≥0.46) and high adjusted CFR (≥2.3%) was observed in 28.9% of counties.

**Conclusions and Relevance:** Social vulnerability is associated with higher COVID-19 case fatality. High social vulnerability and CFR coexist in more than 1 in 4 U.S. counties. These counties should be targeted by public policy interventions to help alleviate the pandemic burden on the most vulnerable population.

**KEY POINTS:** *Question:* Is county-level social vulnerability to disasters associated with the case fatality rate (CFR) and incidence of SARS-CoV-2 infection during the COVID-19 pandemic in the U.S.?

*Findings:* Each unit increase in county-level social vulnerability, measured using the Social Vulnerability Index (SVI), was associated with a 63% higher CFR after adjusting for age and comorbidities. Both CFR and incidence of COVID-19 were significantly higher in counties with lower socio-economic status and higher proportion of minority populations.

*Meaning:* U.S. counties with higher social vulnerability are experiencing greater mortality rates during the COVID-19 pandemic.

## Introduction

Community-level social disadvantage and vulnerability to disasters can influence the incidence of COVID-19 and its adverse outcomes in several ways. For example, lower socioeconomic status (SES) is associated with poor healthcare access, which may increase risk for adverse outcomes.^1^ Labor inequalities, lack of workplace protections, and household overcrowding may decrease the ability to adhere to social-distancing guidelines.^2^ Additionally, race-ethnic minorities and immigrants are less likely to have access to appropriate and timely healthcare.^3^ Evidence suggests that these inequalities contributed to disease spread and severity during the H1N1 influenza pandemic.^4^

Real-time evaluation of the impact of community-level social vulnerability on disease incidence and adverse outcomes during the ongoing COVID-19 pandemic is important to guide public health policy and healthcare resource allocation in the U.S. The Social Vulnerability Index (SVI), created and maintained by the Geospatial Research, Analysis, and Services Program (GRASP) at the Centers for Disease Control and Prevention (CDC) and Agency for Toxic Substances and Disease Registry, is a percentile-based index of county-level vulnerability to disasters.^5,6^ Herein, we report the association of SVI with COVID-19 case-fatality rates (CFR) and incidence in the U.S.

## Methods

County-level data on COVID-19 CFR (deaths per 100 confirmed COVID-19 cases) and incidence (cases per 1000 population) for U.S. counties with at least 50 cases (n=433) were obtained from the Johns Hopkins Center for Systems Science and Engineering database on April 4^th^, 2020.^7^ County-level SVI data for 2018 were obtained from the CDC GRASP database.^8^ As a proxy for county-level medical comorbidity, we utilized Hierarchical Condition Category (HCC) risk scores acquired from the Centers for Medicare and Medicaid Services (CMS), which are based on medical risk profiles and demographics of county Medicare beneficiaries.^9,10^

Generalized linear mixed models, with negative binomial distribution or Poisson distribution when appropriate,^11^ were used to examine the association of outcomes with SVI (reported as percentile of social vulnerability, with higher numbers representing increased vulnerability) and its sub-components including socioeconomic status, household composition, minority status, and housing type/transportation accessibility (**Supplement**).^5,6^ Given differences in COVID-19 testing by state, state-specific random intercepts were incorporated in models to account for correlations among counties within the same state. Covariates included percentage of population aged ≥65 years and average HCC score. A sensitivity analysis was conducted using data from six states (New York, New Jersey, Washington, Massachusetts, Vermont, and Louisiana) with the highest levels of testing as of April 4^th^, 2020.^12^ Age- and HCC score-adjusted CFR and incidence were compared across medians of overall SVI using one-way ANOVA. Lastly, we identified counties with high SVI (≥median) and high adjusted CFR (≥median) as potential targets for public policy interventions. Statistical analyses were performed using R version 3.6.1 (R Foundation for Statistical Computing, Vienna, Austria).

## Results

Of 3,142 counties in the U.S., 433 had >50 COVID-19 confirmed cases, accounting for 283,256 cases and 6,644 deaths as of April 4^th^, 2020. The median SVI was 0.46 [Range: 0.01-1.0], CFR was 1.9% [Range: 0-13.3], and incidence was 1.2 per 1000 people [Range: 0.6-38.8].

There was a significant association between overall SVI and CFR (RR: 1.19 [1.05, 1.34], p=0.005 per-1 unit increase in SVI) that was strengthened after adjustment for percentage of population aged ≥65 years and comorbidities using the county HCC score (RR: 1.63 [1.38, 1.91], p<0.001) (**Table 1**). Sensitivity analysis of six states with the highest levels of testing (n=99 counties, cases=173,612, deaths=4,122) confirmed strong associations between overall SVI and the CFR (RR: 2.58 [2.09, 3.18], p<0.001). In the overall study sample, adjusted CFR was significantly higher in counties with greater vulnerability (SVI<median: 2.18±0.90 vs. ≥median: 2.63±0.99; p<0.001), **Figure 1**. Of the SVI sub-components, socioeconomic status was associated with 2.6-fold, minority status/language with 1.6-fold, and housing type/transport accessibility with 1.9-fold higher CFR in adjusted models (**Table 1**).

**Table 1.** Association of Social Vulnerability Index and its sub-components with case fatality rate and incidence of COVID-19 in the U.S. Risk ratio per-1 unit increase in respective score (indicating greater social vulnerability) is shown

|  | Model 1 |  | Model 2 |  | Model 3 |  |
| --- | --- | --- | --- | --- | --- | --- |
|  | Risk Ratio<br>(95% CI) | P-value | Risk Ratio<br>(95% CI) | P-value | Risk Ratio<br>(95% CI) | P-value |
| **Case Fatality Rate** |  |  |  |  |  |  |
| **Overall Social Vulnerability Index** | 1.19 (1.05, 1.34) | **0.005** | - | - | 1.63 (1.38, 1.91) | **<0.001** |
| Socioeconomic Status | 1.56 (1.37, 1.77) | **<0.001** | 1.73 (1.51, 1.98) | **<0.001** | 2.59 (2.20, 3.07) | **<0.001** |
| Minority Status & Language | 1.08 (0.86, 1.36) | 0.52 | 1.39 (1.07, 1.82) | **0.01** | 1.55 (1.18, 2.07) | **0.002** |
| Housing Type & Transport | 1.44 (1.28, 1.62) | **<0.001** | 1.65 (1.46, 1.88) | **<0.001** | 1.86 (1.63, 2.14) | **<0.001** |
| Household Composition & Disability | 0.83 (0.72, 0.96) | **0.01** | - | - | 0.88 (0.74, 1.01) | 0.08 |
| **Incidence** |  |  |  |  |  |  |
| **Overall Social Vulnerability Index** | 1.30 (0.96, 1.77) | 0.09 | - | - | 1.30 (0.95, 1.78) | 0.21 |
| Socioeconomic Status | 1.50 (1.08, 2.09) | **0.01** | 1.54 (1.11, 2.15) | **0.01** | 1.60 (1.17, 2.20) | **0.03** |
| Minority Status & Language | 1.68 (1.11, 2.53) | **0.01** | 2.04 (1.30, 3.18) | **0.02** | 1.97 (1.44, 2.70) | **0.006** |
| Housing Type & Transport | 1.26 (0.95, 1.66) | 0.11 | 1.30 (0.99, 1.72) | 0.07 | 1.24 (0.90, 1.70) | 0.18 |
| Household Composition & Disability | 0.97 (0.71, 1.33) | 0.85 | - | - | 0.88 (0.65, 1.21) | 0.50 |

Model 1: Unadjusted generalized linear mixed-effects model accounting for state-level clustering.
Model 2: Model 1+ adjusted for percentage of population age $\geq 65$ ; percentage age $\geq 65$ not included as covariates for models for overall Social Vulnerability Index and Household Composition because these indices contain this variable.
Model 3: Model 2+ CMS average Hierarchical Condition Category (HCC) score (proxy for comorbidity)

**Figure 1.**
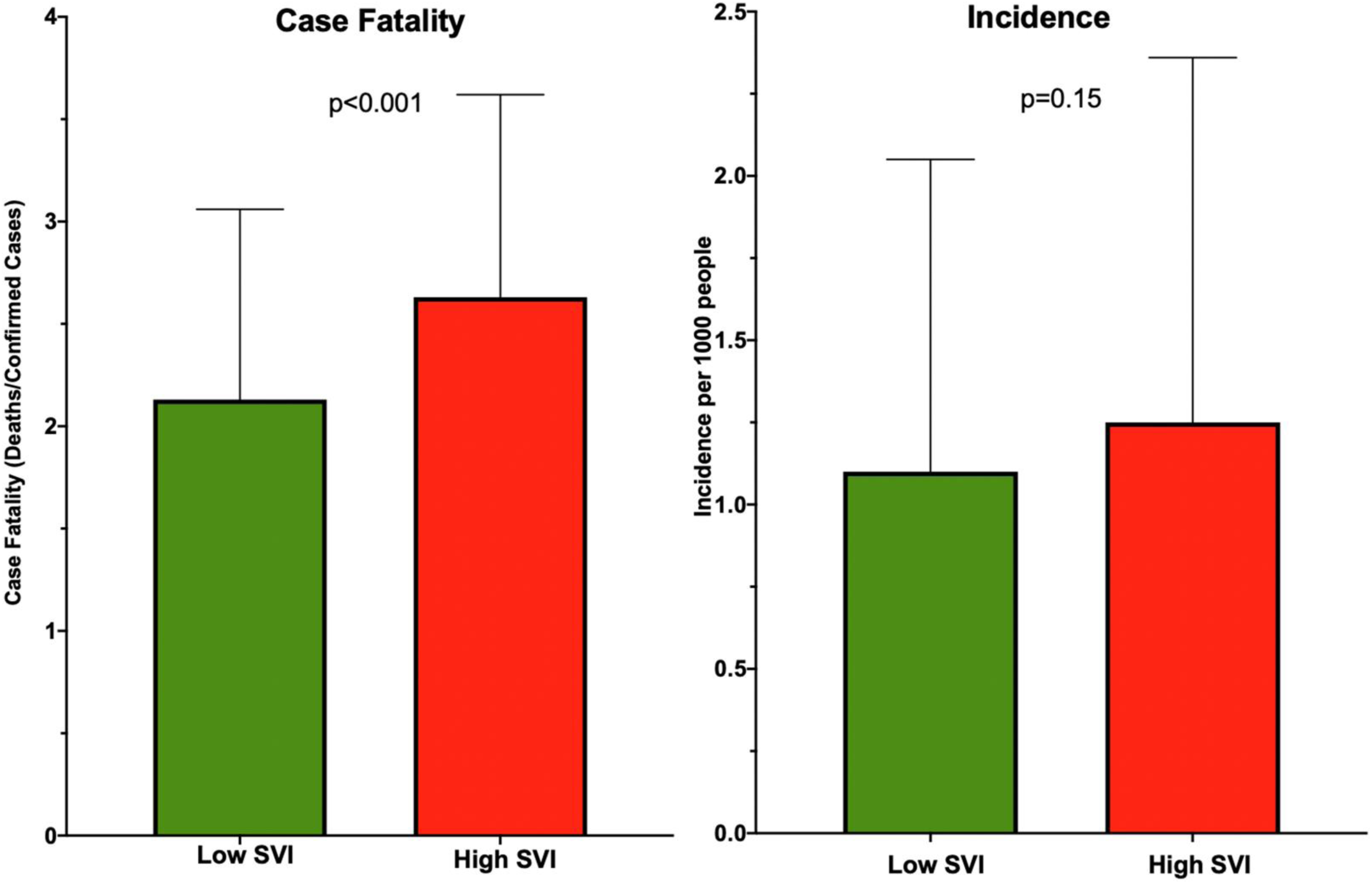
Incidence and Case Fatality Rates of COVID-19 by high (≥median) and low (<median) overall Social Vulnerability Index (SVI) after adjusting for percentage of population age ≥65 years and CMS average Hierarchical Condition Category (HCC) score (proxy for comorbidity).

There was a nominal association between overall SVI and the incidence (RR: 1.30 [0.96, 1.77], p=0.09) that became insignificant after covariate adjustment **(Table 1)**. Thus, the adjusted incidence was similar in counties stratified by the overall SVI (<median:1.10±0.95 vs. ≥median: 1.25±1.11; p=0.15), **Figure 1**. However, two sub-components of the SVI, including socioeconomic status (1.6-fold) and minority status/language (2-fold) were associated with higher incidence in adjusted models (**Table 1**).

In the overall study sample, a combination of high SVI (≥0.46) and high adjusted CFR (≥2.3%) was observed in 125 counties (28.9%, **Table S1**), while 124 counties (28.6%) had SVI and adjusted CFR below respective medians (**Figure 2)**.

**Figure 2.**
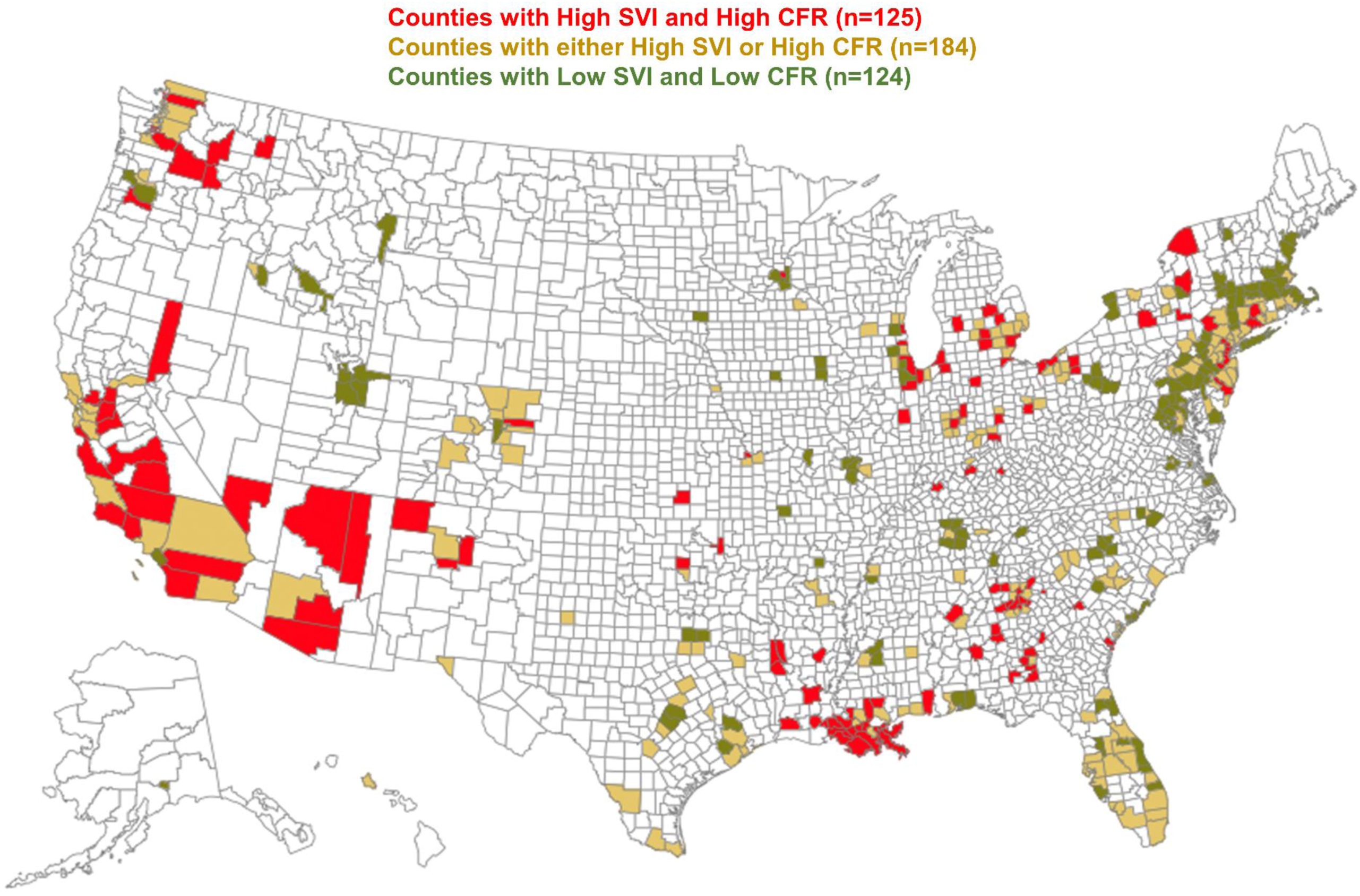
U.S. Map depicting counties (n=433) stratified by median SVI and adjusted CFR. High SVI and adjusted CFR observed in 125 counties, high SVI or adjusted CFR observed in 184 counties, and low SVI and adjusted CFR observed in 124 counties.

## Discussion

Using county-level data and after accounting for state-level clustering, age, and comorbidities, we show that greater social vulnerability is associated with higher COVID-19-related CFR during the first several weeks of the ongoing pandemic in the U.S. This association is driven by lower socioeconomic status, higher minority population prevalence, and poor housing conditions and transport accessibility in these counties. Although the overall SVI was not associated with COVID-19 incidence, it is evident that the incidence is higher in counties with lower socioeconomic status and with greater proportion of minority population.

Our findings are supported by the conceptual model of Blumenshine and colleagues which posits that differences in social position can affect outcomes during pandemics.^13^ Social vulnerability, captured here as socioeconomic status, proportion of minority population, and poor housing conditions, contribute to disparities in both exposure and treatment access, which synergistically contribute to adverse outcomes. Despite earlier calls to address social vulnerabilities in order to reduce adverse outcomes proactively, these disparities were evident during the 2009 H1N1 pandemic,^4,13^ and more than a decade later, continue to account for worse outcomes in vulnerable communities during the COVID-19 pandemic.

More than a quarter of the counties studied, that are located in 22 states with county-level clustering in Louisiana, Georgia, Michigan, Illinois, and California (**Table S1**), had both high social vulnerability and high adjusted CFR. Given the direct association of SVI with CFR, these counties represent COVID-19 ‘hot-spots’ where healthcare resource allocation is urgently needed. Unaddressed, an ensuing “disease-driven poverty trap”, a vicious cycle of disease and worsening social disparity, could have devastating effects in these counties.^14^

A major limitation of the study is our inability to account for the contribution of county-level COVID-19 testing. However, in sensitivity analyses that included states with the highest level of testing, we observed similar associations. In addition, we have only included 433 (13.8%) U.S. counties in our report, but these counties represent the breadth of existing social vulnerability.

This important and timely report demonstrates associations between social vulnerability to disaster and adverse outcomes in the evolving stages of the COVID-19 pandemic in the U.S. Our findings can help guide public policy interventions and resource allocations to improve outcomes in vulnerable communities.

## Data Availability

All data used in the analysis of this manuscript is publicly available.

https://www.cms.gov/Research-Statistics-Data-and-Systems/Statistics-Trends-and-Reports/Medicare-Geographic-Variation/GV_PUF

https://coronavirus.jhu.edu/

https://svi.cdc.gov/

https://covidtracking.com/

## ACKNOWLEDGEMENTS

None

## SUPPLEMENTAL MATERIAL

### Social Vulnerability Index

The social vulnerability index (SVI) is a measure of community resilience to stresses on human health such as disease outbreaks and natural or human-caused disasters.^5,6^ The SVI database and mapping tool has been created by the Geospatial Research, Analysis, and Services Program (GRASP) at the Centers for Disease Control and Prevention (CDC)/Agency for Toxic Substances and Disease Registry. The index helps public health officials and emergency response planners to identify and map communities that are likely to need support before, during, and after a disaster. The SVI uses statistical data from the U.S. Census on 15 variables to determine social vulnerability. These 15 variables are grouped together into four related themes as described below. Each of these variables are ranked from highest to lowest vulnerability across census tracts in the U.S. and a percentile rank is calculated for each variable, theme, and the overall SVI. We have used the 2018 SVI data at the county level in the current analysis.^3^

|  |  |
| --- | --- |
| **Socioeconomic Status** | Below Poverty<br>Unemployed<br>Income<br>No High School Diploma |
| **Household composition and Disability** | Age 65 years or older<br>Age 17 years or younger<br>Older than Age 5 years with a Disability |
| **Minority Status and Language** | Minority<br>Speak English "Less than Well" |
| **Housing Type and Transportation** | Multi-Unit Structures<br>Mobile Homes<br>Crowding<br>No Vehicle<br>Group Quarters |

**Table S1.** Counties with high Social Vulnerability Index (≥median, 0.46) and high Adjusted Case Fatality Rate (≥median, 2.3%)

| State | County | Area<br>(Square<br>miles) | Total<br>population | Adjusted<br>Incidence<br>per 1000<br>population | Adjusted<br>Case<br>Fatality<br>Rate | Social<br>Vulnerability<br>Index |
| --- | --- | --- | --- | --- | --- | --- |
| ALABAMA | Chambers | 596.56 | 33826 | 0.77 | 2.99 | 0.74 |
| ALABAMA | Jefferson | 1111.38 | 659892 | 0.75 | 2.88 | 0.66 |
| ALABAMA | Lee | 607.55 | 159287 | 0.75 | 3.20 | 0.66 |
| ALABAMA | Mobile | 1229.37 | 414659 | 0.77 | 2.87 | 0.75 |
| ALABAMA | Montgomery | 785.30 | 226941 | 0.79 | 3.14 | 0.84 |
| ARIZONA | Coconino | 18618.67 | 140217 | 0.88 | 2.70 | 0.71 |
| ARIZONA | Navajo | 9949.88 | 108705 | 0.95 | 2.67 | 1.00 |
| ARIZONA | Pima | 9187.22 | 1019722 | 0.92 | 2.72 | 0.88 |
| ARIZONA | Pinal | 5365.84 | 419721 | 0.92 | 2.42 | 0.86 |
| CALIFORNIA | Fresno | 5958.10 | 978130 | 0.35 | 2.76 | 0.96 |
| CALIFORNIA | Kern | 8132.26 | 883053 | 0.35 | 2.49 | 0.97 |
| CALIFORNIA | Monterey | 3281.72 | 433212 | 0.33 | 3.16 | 0.80 |
| CALIFORNIA | Riverside | 7206.56 | 2383286 | 0.33 | 2.51 | 0.79 |
| CALIFORNIA | Sacramento | 965.44 | 1510023 | 0.33 | 2.44 | 0.73 |
| CALIFORNIA | San Diego | 4208.76 | 3302833 | 0.31 | 2.41 | 0.56 |
| CALIFORNIA | San Joaquin | 1392.39 | 732212 | 0.34 | 2.71 | 0.89 |
| CALIFORNIA | Santa<br>Barbara | 2735.14 | 443738 | 0.33 | 2.98 | 0.71 |
| CALIFORNIA | Santa Cruz | 445.12 | 273765 | 0.31 | 3.05 | 0.54 |
| CALIFORNIA | Solano | 821.75 | 438530 | 0.32 | 2.53 | 0.59 |
| CALIFORNIA | Stanislaus | 1496.02 | 539301 | 0.34 | 2.60 | 0.87 |
| CALIFORNIA | Tulare | 4824.38 | 460477 | 0.35 | 2.65 | 0.94 |
| CALIFORNIA | Ventura | 1842.47 | 848112 | 0.31 | 2.40 | 0.48 |
| COLORADO | Adams | 1166.26 | 497115 | 1.66 | 2.51 | 0.59 |
| CONNECTICUT | Hartford | 734.96 | 894730 | 1.05 | 3.03 | 0.52 |
| CONNECTICUT | New Haven | 604.52 | 859339 | 1.06 | 3.00 | 0.52 |
| GEORGIA | Bartow | 459.52 | 103620 | 1.42 | 3.27 | 0.59 |
| GEORGIA | Carroll | 499.03 | 116022 | 1.49 | 4.05 | 0.78 |
| GEORGIA | Chatham | 433.12 | 287049 | 1.46 | 3.88 | 0.69 |
| GEORGIA | Clarke | 119.22 | 124602 | 1.49 | 3.96 | 0.77 |
| GEORGIA | Clayton | 141.67 | 278666 | 1.55 | 3.08 | 0.91 |
| GEORGIA | DeKalb | 267.73 | 743187 | 1.45 | 3.38 | 0.66 |
| GEORGIA | Dougherty | 328.64 | 91049 | 1.56 | 3.55 | 0.95 |
| GEORGIA | Douglas | 200.10 | 141840 | 1.40 | 2.95 | 0.54 |
| GEORGIA | Early | 512.60 | 10348 | 1.55 | 4.26 | 0.92 |
| GEORGIA | Floyd | 509.82 | 96824 | 1.54 | 3.83 | 0.89 |
| GEORGIA | Fulton | 526.74 | 1021902 | 1.40 | 3.36 | 0.53 |
| GEORGIA | Hall | 392.98 | 195961 | 1.45 | 3.82 | 0.66 |
| GEORGIA | Houston | 376.06 | 151682 | 1.41 | 3.23 | 0.57 |
| GEORGIA | Mitchell | 512.16 | 22432 | 1.58 | 3.62 | 0.99 |
| GEORGIA | Richmond | 324.32 | 201463 | 1.55 | 3.98 | 0.93 |
| GEORGIA | Rockdale | 129.81 | 89011 | 1.41 | 3.51 | 0.55 |
| GEORGIA | Sumter | 482.87 | 30352 | 1.57 | 3.44 | 0.98 |
| GEORGIA | Terrell | 335.74 | 8859 | 1.56 | 3.62 | 0.96 |
| ILLINOIS | Champaign | 996.11 | 209448 | 0.68 | 2.46 | 0.50 |
| ILLINOIS | Cook | 944.93 | 5223719 | 0.71 | 2.34 | 0.68 |
| ILLINOIS | Kankakee | 676.50 | 111061 | 0.71 | 2.40 | 0.67 |
| INDIANA | Allen | 657.32 | 370016 | 1.05 | 2.79 | 0.52 |
| INDIANA | Lake | 498.82 | 486849 | 1.11 | 2.61 | 0.73 |
| INDIANA | Madison | 451.92 | 129505 | 1.10 | 2.70 | 0.67 |
| INDIANA | Marion | 396.44 | 944523 | 1.13 | 2.74 | 0.80 |
| INDIANA | St. Joseph | 457.89 | 269240 | 1.08 | 3.11 | 0.62 |
| KANSAS | Sedgwick | 997.48 | 512064 | 0.53 | 3.26 | 0.54 |
| KANSAS | Wyandotte | 151.64 | 164345 | 0.59 | 2.70 | 0.92 |
| KENTUCKY | Daviess | 458.39 | 99937 | 0.53 | 3.56 | 0.62 |
| KENTUCKY | Fayette | 283.64 | 318734 | 0.51 | 3.61 | 0.49 |
| KENTUCKY | Jefferson | 380.69 | 767154 | 0.52 | 3.43 | 0.57 |
| LOUISIANA | Acadia | 655.17 | 62568 | 2.80 | 3.66 | 0.92 |
| LOUISIANA | Assumption | 338.62 | 22714 | 2.70 | 3.52 | 0.78 |
| LOUISIANA | Bossier | 840.30 | 126131 | 2.57 | 3.46 | 0.59 |
| LOUISIANA | Caddo | 878.83 | 248361 | 2.75 | 3.46 | 0.84 |
| LOUISIANA | Calcasieu | 1064.29 | 200182 | 2.67 | 3.96 | 0.74 |
| LOUISIANA | De Soto | 876.40 | 27216 | 2.78 | 3.78 | 0.89 |
| LOUISIANA | East Baton Rouge | 455.34 | 444094 | 2.62 | 3.18 | 0.67 |
| LOUISIANA | Iberia | 574.09 | 72691 | 2.84 | 4.21 | 0.96 |
| LOUISIANA | Iberville | 618.63 | 32956 | 2.78 | 3.19 | 0.89 |
| LOUISIANA | Jefferson | 295.66 | 435300 | 2.59 | 3.15 | 0.61 |
| LOUISIANA | Lafayette | 268.74 | 240091 | 2.59 | 3.68 | 0.62 |
| LOUISIANA | Lafourche | 1068.36 | 98214 | 2.69 | 3.89 | 0.77 |
| LOUISIANA | Orleans | 169.43 | 389648 | 2.68 | 3.11 | 0.74 |
| LOUISIANA | Ouachita | 610.40 | 156075 | 2.76 | 3.71 | 0.85 |
| LOUISIANA | Plaquemines | 780.34 | 23373 | 2.65 | 3.50 | 0.71 |
| LOUISIANA | Rapides | 1320.34 | 131546 | 2.80 | 4.24 | 0.91 |
| LOUISIANA | St. Bernard | 377.51 | 45694 | 2.70 | 3.46 | 0.77 |
| LOUISIANA | St. James | 241.54 | 21357 | 2.60 | 3.38 | 0.63 |
| LOUISIANA | St. John the Baptist | 214.32 | 43446 | 2.68 | 3.26 | 0.75 |
| LOUISIANA | St. Martin | 737.22 | 53752 | 2.71 | 4.11 | 0.78 |
| LOUISIANA | St. Mary | 555.58 | 51734 | 2.80 | 4.38 | 0.92 |
| LOUISIANA | Tangipahoa | 791.31 | 130504 | 2.78 | 3.50 | 0.89 |
| LOUISIANA | Terrebonne | 1229.83 | 112587 | 2.79 | 4.08 | 0.90 |
| LOUISIANA | Washington | 669.53 | 46457 | 2.86 | 3.26 | 1.00 |
| MICHIGAN | Berrien | 567.78 | 154807 | 1.10 | 5.50 | 0.65 |
| MICHIGAN | Genesee | 636.96 | 409361 | 1.11 | 3.91 | 0.66 |
| MICHIGAN | Ingham | 556.13 | 289564 | 1.08 | 4.56 | 0.57 |
| MICHIGAN | Jackson | 701.88 | 158913 | 1.07 | 4.51 | 0.55 |
| MICHIGAN | Kent | 848.05 | 643140 | 1.05 | 4.22 | 0.48 |
| MICHIGAN | Saginaw | 800.47 | 192778 | 1.12 | 4.24 | 0.70 |
| MICHIGAN | Wayne | 611.99 | 1761382 | 1.17 | 3.89 | 0.87 |
| MINNESOTA | Ramsey | 152.22 | 541493 | 0.35 | 2.63 | 0.63 |
| NEVADA | Clark | 7891.65 | 2141574 | 0.69 | 2.63 | 0.74 |
| NEVADA | Washoe | 6301.42 | 450486 | 0.65 | 2.84 | 0.53 |
| NEW JERSEY | Atlantic | 555.51 | 268539 | 2.82 | 3.27 | 0.79 |
| NEW JERSEY | Camden | 221.35 | 507367 | 2.69 | 2.91 | 0.61 |
| NEW JERSEY | Essex | 126.08 | 793555 | 2.86 | 2.79 | 0.84 |
| NEW JERSEY | Hudson | 46.19 | 668631 | 2.71 | 2.61 | 0.64 |
| NEW JERSEY | Mercer | 224.43 | 368762 | 2.65 | 2.75 | 0.56 |
| NEW JERSEY | Middlesex | 309.22 | 826698 | 2.59 | 2.83 | 0.47 |
| NEW JERSEY | Passaic | 186.01 | 504041 | 2.81 | 3.20 | 0.78 |
| NEW JERSEY | Union | 102.77 | 553066 | 2.70 | 2.92 | 0.62 |
| NEW MEXICO | Bernalillo | 1161.26 | 677692 | 0.49 | 2.34 | 0.68 |
| NEW MEXICO | San Juan | 5517.20 | 127455 | 0.52 | 2.44 | 0.92 |
| NEW MEXICO | Santa Fe | 1910.10 | 148917 | 0.47 | 2.68 | 0.53 |
| NEW YORK | Broome | 705.82 | 194402 | 4.33 | 2.40 | 0.62 |
| NEW YORK | Oneida | 1212.33 | 230782 | 4.43 | 2.38 | 0.69 |
| NEW YORK | St.<br>Lawrence | 2679.27 | 109558 | 4.49 | 2.67 | 0.75 |
| NEW YORK | Steuben | 1390.55 | 96927 | 4.17 | 2.30 | 0.47 |
| NEW YORK | Sullivan | 968.14 | 75211 | 4.54 | 2.38 | 0.79 |
| OHIO | Cuyahoga | 457.18 | 1253783 | 0.48 | 2.69 | 0.68 |
| OHIO | Franklin | 532.41 | 1275333 | 0.46 | 2.55 | 0.51 |
| OHIO | Hamilton | 405.95 | 812037 | 0.46 | 2.58 | 0.54 |
| OHIO | Lorain | 491.21 | 306713 | 0.46 | 2.51 | 0.51 |
| OHIO | Lucas | 340.71 | 432379 | 0.48 | 2.74 | 0.69 |
| OHIO | Mahoning | 411.51 | 231064 | 0.47 | 2.73 | 0.59 |
| OHIO | Montgomery | 461.49 | 532034 | 0.48 | 2.64 | 0.67 |
| OHIO | Trumbull | 618.04 | 201794 | 0.45 | 2.65 | 0.46 |
| OKLAHOMA | Oklahoma | 708.82 | 782051 | 0.53 | 3.59 | 0.68 |
| OKLAHOMA | Tulsa | 570.32 | 642781 | 0.52 | 3.70 | 0.62 |
| OREGON | Marion | 1180.55 | 335553 | 0.44 | 2.48 | 0.85 |
| WASHINGTON | Benton | 1699.98 | 194168 | 0.91 | 4.92 | 0.51 |
| WASHINGTON | Grant | 2679.49 | 94860 | 0.99 | 5.79 | 0.85 |
| WASHINGTON | Pierce | 1667.81 | 859840 | 0.90 | 4.86 | 0.49 |
| WASHINGTON | Skagit | 1730.22 | 123907 | 0.91 | 5.71 | 0.54 |
| WASHINGTON | Spokane | 1763.89 | 497875 | 0.90 | 4.92 | 0.48 |
| WASHINGTON | Yakima | 4294.52 | 249325 | 1.01 | 6.31 | 0.93 |
| WISCONSIN | Kenosha | 271.86 | 168330 | 0.61 | 2.78 | 0.51 |
| WISCONSIN | Milwaukee | 241.48 | 954209 | 0.66 | 2.44 | 0.81 |

